# Elevated FGF23 levels predict cardiovascular and cerebrovascular events in acute ischaemic stroke patients

**DOI:** 10.1101/2024.12.12.24318962

**Authors:** Lingyun Gu, Liping Shen, Weizhang Li, Jinfeng Zhou, Qi Jin, Jiangsheng Yang, Junyou Cui, Zufu Zhu, Zhuowen Xu, Yulong Weng

## Abstract

**BACKGROUND:** There have been recent studies on fibroblast growth factor 23 (FGF23) and acute ischaemic stroke (AIS), but some of the results were conflicting. Therefore, the objective of this study was to assess whether baseline FGF23 was associated with the occurrence of major adverse cardiovascular and cerebrovascular events (MACCE) in AIS patients.

**METHODS:** This study enrolled 394 patients with AIS and followed for a median period of 43 months. 130 people who underwent health check-ups in our hospital were selected as the control group. The study endpoint was the first occurrence of MACCE outside the hospital. Serum FGF23 levels were quantified using an enzyme-linked immunosorbent assay.

**RESULTS:** Serum FGF23 levels were found to be significantly higher in the AIS group than in the control group (525.14±167.40 vs 338.62±161.71, *P*<0.05). Furthermore, serum FGF23 levels were observed to be elevated in the MACCE group in comparison to the no MACCE group (646.09±164.05 vs 491.87±152.54, *P*<0.05). According to the receiver operating characteristic curve analysis, the predictive value of the combination of the three [FGF23, National Institutes of Health Stroke Scale (NIHSS) and modified Rankin Scale (mRS)] for the occurrence of MACCE in AIS was higher than that of any of them (*P*<0.05), while the difference between the three was not statistically significant. The cumulative MACCE-free survival was found to be significantly higher in the group with low FGF23 levels than in the group with high FGF23 levels, as demonstrated by Kaplan-Meier analysis (*P*<0.05). The multifactorial Cox analysis revealed that the elevated baseline serum FGF23 level (HR: 3.731, 95% CI: 2.157-6.452) was a significant predictor of MACCE in AIS patients.

**CONCLUSIONS:** Elevated baseline serum FGF23 levels were considered to be a valid predictor of MACCE in patients with AIS.

---

Acute ischaemic stroke (AIS), as one of the common clinical neurological disorders, is the second leading cause of disability and death worldwide.^1^ AIS patients have a high risk of major adverse cardiovascular and cerebrovascular events (MACCE), resulting in a significant socioeconomic burden.^2–4^ The study found that the cumulative risk of recurrence of thrombotic arterial events, including the recurrent AIS, cardiovascular events and death, was greater than 11 percent within five years of the first occurrence of AIS.^5^ The risk of AIS recurrence is about 15 percent 10 years after AIS onset, and the risk of hospitalisation for cardiovascular events and AIS recurrence is about 20 percent 15 years later.^6–8^ Therefore, early identification and aggressive treatment of AIS patients at high risk of developing MACCE is clinically important.

Although multiple methods can predict the prognosis of patients with AIS, the predictive efficacy is limited. However, biomarkers may provide additional information to existing predictive methods to guide clinical practice.^9^ It has been found that several biomarkers, such as osteoprotegerin, C-reactive protein, galectin-3, and brain-derived neurotrophic factor, have been associated with the prognosis of AIS and have good predictive value.^10–13^ In recent years, studies related to fibroblast growth factor 23 (FGF23) in cerebrovascular disease have received increasing attention.

FGF23 is a phosphotropic hormone secreted mainly by osteoblasts and osteoclasts and acts in various organs via endocrine forms.^14^ FGF23 in the blood could be transported into the brain through the blood-brain barrier or the blood-cerebrospinal fluid barrier.^15^ Previous studies have found that FGF23 levels were associated with the severity and prognosis of a variety of diseases, including diabetes mellitus, chronic coronary heart disease, heart failure and chronic kidney disease.^16–19^

Recent studies found that serum FGF23 levels were significantly elevated in patients with AIS and correlated with short-term prognosis.^20^ The North Manhattan study found that elevated FGF23 levels were a risk factor for stroke and cerebral haemorrhage events in the population and were associated with small vessel disease and cerebral infarction, but not with chronic kidney disease.^21,22^ A meta-analysis of seven high-quality prospective studies found a significant positive association between baseline FGF23 levels and stroke incidence, and that elevated FGF23 levels were an independent predictor of stroke.^23^

However, there are still inconsistencies in the findings of studies correlating FGF23 levels with AIS. The Multi-Ethnic Atherosclerosis Study (MESA) that included 6,547 members of the general population initially free of cardiovascular disease found that FGF23 levels were not associated with stroke.^24^ The Reasons for Geographic and Racial Differences in Stroke (REGARDS) study including 615 stroke patients found that elevated levels of FGF23 were associated with cardioembolic stroke, but not with other stroke subtypes, in community-living adults.^25^ Moreover, there are fewer studies correlating FGF23 levels with the occurrence of MACCE in AIS. Therefore, the purpose of this study was to assess whether baseline FGF23 levels were associated with AIS and had the ability to predict the occurrence of MACCE.

## METHODS

### Study population

This was a prospective study. This study enrolled 394 patients who met the definition of AIS in the Chinese AIS Diagnostic and Treatment Guidelines for AIS.^26^ All these patients received standardised treatment according to the guidelines between January 2020 and September 2020 at the affiliated jiangyin clinical college of xuzhou medical university. 130 cases of the general population who had physical examination during the same period served as the control group. The following were the exclusion criteria: recurrent ischemic stroke or cerebral haemorrhage with neurodegenerative diseases such as Parkinson’s disease, Alzheimer’s disease or vascular dementia, transient ischemic attack, haematological diseases, autoimmune diseases, co-infections, inability to follow up. This study was obtained approval from the Ethics Committee of Jiangyin People’s Hospital (Approval No. 2020008). Informed consent was obtained from the patients or their families (patients who were unable to communicate) prior to enrolment of all patients.

### Clinical and Laboratory Assessments

After the patients were enrolled, a detailed neurological physical examination was performed and recorded after detailed questioning of their current and past medical history by experts with extensive clinical experience. The height and weight of the patients were recorded to calculate the body mass index (BMI). The National Institutes of Health Stroke Scale (NIHSS) score was used to assess the severity of AIS immediately after patient enrolment.^27^ The Oxfordshire Community Stroke Project criteria and the TOAST (Trialof Org 10172 in Acute Stroke Treatment) classification criteria were used to identify stroke syndromes and stroke etiology in the enrolled patients, respectively.^28^

Elbow venous blood was drawn from fasting enrolled patients on the morning of the second day of hospitalisation. A portion of the blood sample was delivered to the central laboratory where total cholesterol, triglycerides, low-density lipoprotein cholesterol (LDL-C), high-density lipoprotein cholesterol (HDL-C), and creatinine levels were measured using the Roche e602 and c701 modules, respectively. The other part of the blood sample was left at room temperature for 1 hour and then centrifuged at 3000 r/min for 15 minutes to obtain the upper serum layer, which was stored at -80°C in a refrigerator. After following the instructions of the human enzyme-linked immunosorbent assay kit (Elabscience), serum FGF23 concentration was measured at 450 nm using an enzyme marker.

### Follow-up

Patients or their families were followed up monthly through clinic visits or by telephone. The median follow-up time in this study as of December 2023 was 43 months (interquartile range, 39-44 months). The endpoint event in this study was the first out-of-hospital MACCE, including AIS, lacunar cerebral infarction, cerebral haemorrhage, non-fatal myocardial infarction, heart failure, and all-cause mortality. At the end of the follow-up, a modified Rankin Scale (mRS) was used to assess the neurological recovery of the enrolled patients.

### Statistical analysis

Statistical analysis was performed using the SPSS 25.0 statistical package. Quantitative variables were expressed as means ± standard deviations and compared using Student’s t-test. Categorical variables were expressed as absolute numbers (percentages) and compared using the chi-squared test and the Mann-Whitney-Wilcoxon test. The receiver operating characteristic (ROC) curve was used to determine the best cut-off value for each variable to predict the occurrence of MACCE in patients with AIS. Comparisons between different ROC curves were Z-tested by MedCalc software. Kaplan-Meier survival curves were used for survival analysis and compared using the log-rank test. Univariate and multivariate cox proportional hazards models were used to analyse the relationships between serum FGF23 levels and the occurrence of MACCE, calculating hazard ratios (HRs) and 95% confidence intervals (CIs). All variables that reached statistical significance in the univariate cox analysis were included in the multivariate cox analysis. A p-value < 0.05 was considered statistically significant (two-tailed).

## RESULTS

### Baseline Characteristics of Study Participants

The comparisons of demographic and baseline clinical characteristics between the AIS and control groups were shown in Table 1. Serum FGF23 levels were significantly higher in the AIS group than in the control group, and the difference was statistically significant (525.14±167.40 vs 338.62±161.71, *P*<0.05). The prevalence of hypertension, diabetes mellitus and atrial fibrillation was significantly higher in the AIS group than in the control group (*P*<0.05). There was also a statistical difference between the two groups in terms of BMI and blood LDL-C levels, whereas there was no statistical difference in terms of gender, age, blood total cholesterol, triglycerides, HDL-C and creatinine levels.

**Table 1.** The Demographic and Baseline Clinical Characteristics of the Patients.

| Variables | AIS group<br>(n=394) | Control group<br>(n=130) | *P* |
| --- | --- | --- | --- |
| Male, n (%) | 236 (59.90%) | 80 (61.54%) | 0.740 |
| Age (years) | 67.32±12.29 | 65.70±11.42 | 0.186 |
| BMI (kg/m<sup>2</sup>) | 23.34±2.25 | 22.50±2.12 | 0.000 |
| Hypertension, n (%) | 98 (24.87%) | 12 (9.23%) | 0.000 |
| Diabetes mellitus, n (%) | 67 (17.01%) | 10 (7.69%) | 0.009 |
| Atrial fibrillation, n (%) | 62 (15.74%) | 6 (4.62%) | 0.001 |
| Total cholesterol (mmol/L) | 4.42±0.95 | 4.28±0.81 | 0.132 |
| Triglyceride (mmol/L) | 1.82±1.33 | 1.86±1.61 | 0.758 |
| LDL-C (mmol/L) | 3.09±0.97 | 2.53±0.74 | 0.000 |
| HDL-C (mmol/L) | 0.96±0.26 | 0.99±0.27 | 0.240 |
| Creatinine (μmol/L) | 78.92±33.72 | 73.92±30.19 | 0.133 |
| FGF23 (pg/ml) | 525.14±167.40 | 338.62±161.71 | 0.000 |
| NIHSS | 14.47±8.20 | - | - |
| mRS | 2.45±1.25 | - | - |
| Stroke syndrome, n (%) |  | - | - |
| TACS | 91 (23.10%) | - | - |
| PACS | 123 (31.22%) | - | - |
| LACS | 70 (17.77%) | - | - |
| POCS | 110 (27.92%) | - | - |
| Stroke etiology, n (%) |  | - | - |
| Large-vessel occlusive | 71 (18.02%) | - | - |
| Small-vessel occlusive | 88 (22.34%) | - | - |
| Cardioembolic | 89 (22.59%) | - | - |
| Multiple causes | 84 (21.32%) | - | - |
| Unknown | 62 (15.74%) | - | - |

AIS, acute ischemic stroke BMI; body mass index; LDL-C, low density lipoprotein cholesterol; HDL-C, high density lipoprotein cholesterol; FGF23, fibroblast growth factor 23; NIHSS, national institute of health stroke scale; mRS, modified rankin scale; TACS, total anterior circulation syndrome; PACS, partial anterior circulation syndrome; LACS, lacunar syndrome; POCS, posterior circulation syndrome.

### Comparison of clinical data between the MACCE and no MACCE groups of AIS patients

In this study, 394 patients with AIS were followed up for a median duration of 43 months (interquartile range, 39-44 months). By the end of follow-up, a total of 85 MACCEs were recorded, including 33 AIS recurrences, 17 lacunar cerebral infarctions, 4 cerebral haemorrhages, 11 non-fatal myocardial infarctions, 6 heart failures and 9 all-cause deaths. Patients with AIS were divided into a MACCE group and a no MACCE group, based on whether or not a MACCE occurred. The comparison of demographic and baseline clinical characteristics between the two groups was shown in Table 2.

**Table 2.** The Comparison of clinical data between the MACCE and no MACCE group in AIS patients.

| Variables | MACCE group<br>(n=85) | NO MACCE group<br>(n=309) | *P* |
| --- | --- | --- | --- |
| Male, n (%) | 53 (62.35%) | 183 (59.22%) | 0.602 |
| Age (years) | 70.95±12.93 | 66.32±11.94 | 0.002 |
| BMI (kg/m<sup>2</sup>) | 23.67±2.43 | 23.25±2.19 | 0.125 |
| Hypertension, n (%) | 35 (41.18%) | 63 (20.39%) | 0.000 |
| Diabetes mellitus, n (%) | 19 (22.35%) | 48 (15.53%) | 0.138 |
| Atrial fibrillation, n (%) | 29 (34.12%) | 33 (10.68%) | 0.000 |
| Total cholesterol (mmol/L) | 4.42±0.90 | 4.42±0.96 | 0.945 |
| Triglyceride (mmol/L) | 1.78±1.08 | 1.83±1.39 | 0.776 |
| LDL-C (mmol/L) | 3.48±0.93 | 2.98±0.96 | 0.000 |
| HDL-C (mmol/L) | 0.90±0.22 | 0.98±0.26 | 0.013 |
| Creatinin (μmol/L) | 80.09±35.69 | 78.60±33.21 | 0.719 |
| FGF23 (pg/ml) | 646.09±164.05 | 491.87±152.54 | 0.000 |
| NIHSS | 21.26±9.14 | 12.60±6.84 | 0.000 |
| mRS | 3.69±1.39 | 2.11±0.96 | 0.000 |
| Stroke syndrome, n (%) |  |  |  |
| TACS | 34 (40.00%) | 57 (18.45%) | 0.000 |
| PACS | 20 (23.53%) | 103 (33.33%) | 0.084 |
| LACS | 12 (14.12%) | 58 (18.77%) | 0.320 |
| POCS | 19 (22.35%) | 91 (29.45%) | 0.196 |
| Stroke etiology, n (%) |  |  |  |
| Large-vessel occlusive | 12 (14.12%) | 59 (19.09%) | 0.290 |
| Small-vessel occlusive | 19 (22.35%) | 69 (22.33%) | 0.996 |
| Cardioembolic | 28 (32.94%) | 61 (19.74%) | 0.010 |
| Multiple causes | 16 (18.82%) | 68 (22.01%) | 0.526 |
| Unknown | 10 (11.76%) | 52 (16.83%) | 0.256 |

MACCE, major adverse cardiovascular and cerebrovascular events; AIS, acute ischemic stroke BMI; body mass index; LDL-C, low density lipoprotein cholesterol; HDL-C, high density lipoprotein cholesterol; FGF23, fibroblast growth factor 23; NIHSS, national institute of health stroke scale; mRS, modified rankin scale; TACS, total anterior circulation syndrome; PACS, partial anterior circulation syndrome; LACS, lacunar syndrome; POCS, posterior circulation syndrome.

Serum FGF23 levels were significantly elevated in the MACCE group compared to the no MACCE group (646.09±164.05 vs 491.87±152.54, *P*<0.05). The MACCE group exhibited superior NIHSS and mRS scores in comparison to the NO MACCE group (*P*<0.05). The prevalence of total anterior circulation syndrome and cardioembolic was markedly elevated in the MACCE group than in the NO MACCE group (*P*<0.05). There was a statistical difference between the two groups in terms of age, history of hypertension, history of atrial fibrillation, blood LDL cholesterol and HDL cholesterol levels, whereas there was no statistically significant difference in terms of gender, BMI, history of diabetes mellitus, blood levels of total cholesterol, triglycerides and creatinine.

### ROC analyses to predict the occurrence of MACCE in AIS patients

To assess the value of FGF23 in predicting the occurrence of MACCE in patients with AIS, a ROC analysis was performed, and the NIHSS and mRS were selected for comparison (Figure 1). According to the ROC analysis, the area under the curve (AUC) of FGF23 was 0.754 (95% CI: 0.693-0.816), which was lower than that of NIHSS (AUC: 0.774, 95% CI: 0.714-0.833) and mRS (AUC: 0.809, 95% CI: 0.751-0.867). However, the difference between the three was not statistically significant. The AUC of the combination of the three was 0.851 (95% CI: 0.797-0.905), which was greater than that of mRS, NIHSS, and FGF23 (Z: 2.557, 3.051, and 3.679, respectively; all *P*<0.05).

**Figure 1.**
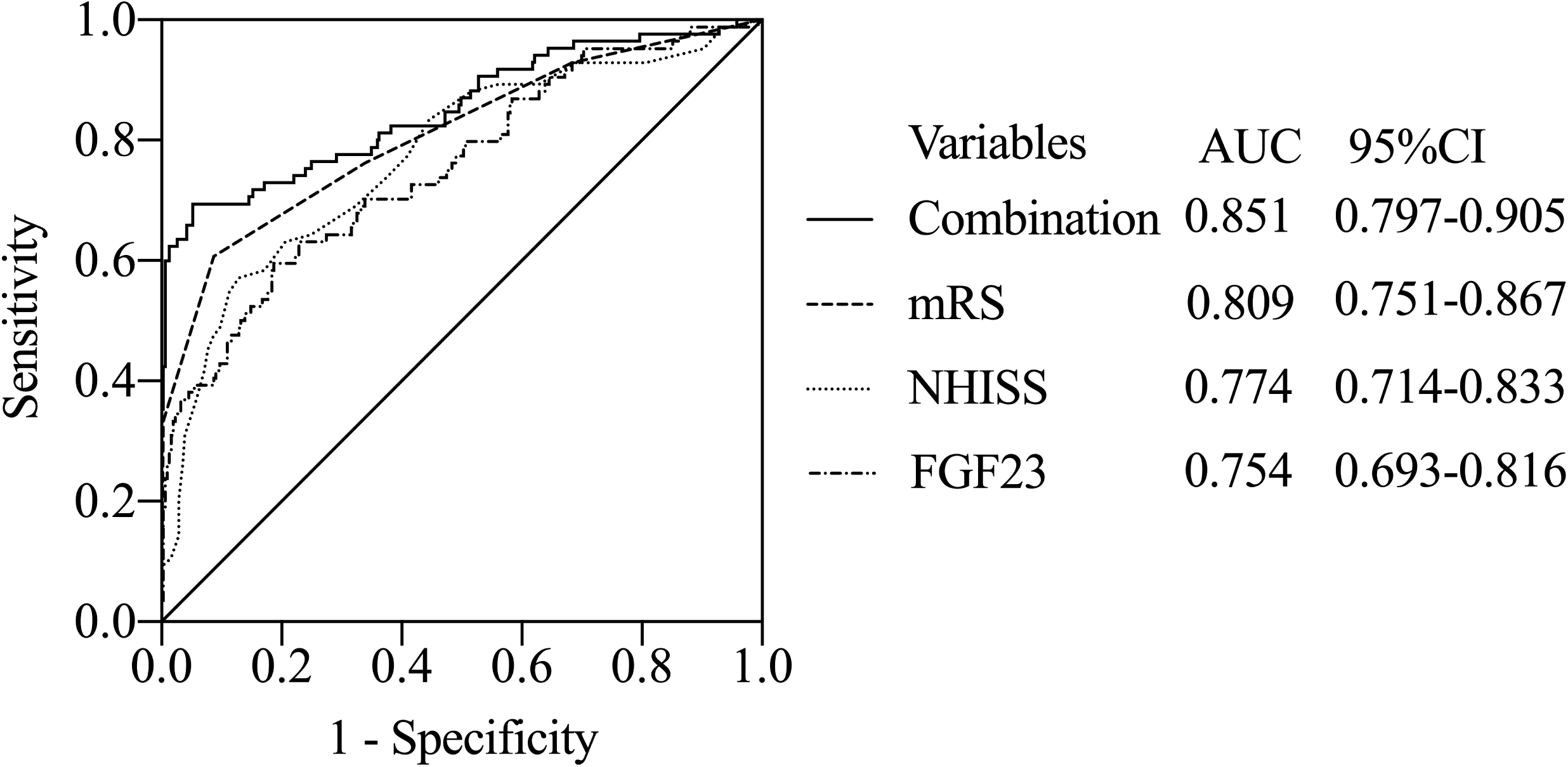
ROC analyses to predict the occurrence of MACCE in AIS patients. AUC, area under the curve; mRS, modified Rankin Scale; NIHSS, national institute of health stroke scale; FGF23, fibroblast growth factor 23.

### Kaplan-meier analysis of FGF23 and the occurrence of MACCE in AIS patients

The optimal cut-off value of FGF23 (642.42pg/mL) based on the Jordon’s index was then used to perform survival analyses. A kaplan-meier analysis based on this cut-off value showed a significant difference between the two groups in terms of cumulative survival rates without MACCE (Figure 2). The cumulative MACCE-free survival rate was significantly higher in the low FGF23 level group than in the high FGF23 level group (*P*<0.05). Moreover, the difference between the two groups gradually increased over time, indicating that high FGF23 levels were associated with lower MACCE-free survival.

**Figure 2.**
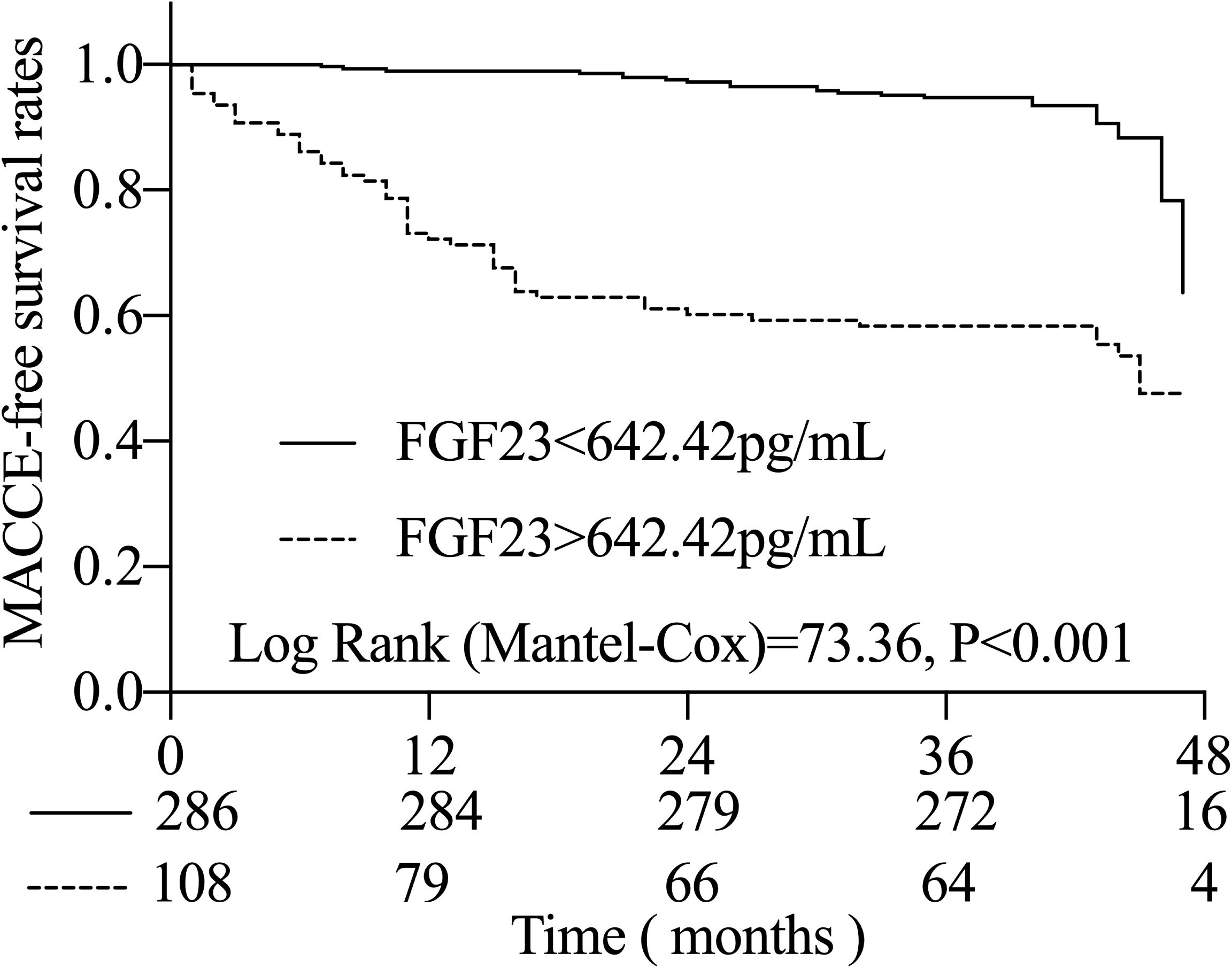
Kaplan-meier analysis of FGF23 and the occurrence of MACCE in AIS patients. MACCE, major adverse cardiovascular and cerebrovascular events; FGF23, fibroblast growth factor 23.

### Univariate and Multivariate COX Analysis for MACCE in AIS Patients

The univariate COX analysis showed that elevated baseline serum FGF23 levels along with advanced age, elevated BMI, previous history of hypertension and atrial fibrillation, elevated LDL-C levels, high NIHSS and mRS scores, and Stroke syndrome were predictive factors for the development of MACCE in AIS patients (Table 3). Further multifactorial COX analyses of statistically significant metrics showed that elevated baseline serum FGF23 levels (HR: 3.731, 95%CI:2.157-6.452) remained a potent predictor of the development of MACCE in patients with AIS, as did advanced age, a history of previous AF, elevated LDL-C levels, high NIHSS and mRS scores, and Stroke syndrome.

**Table 3.**
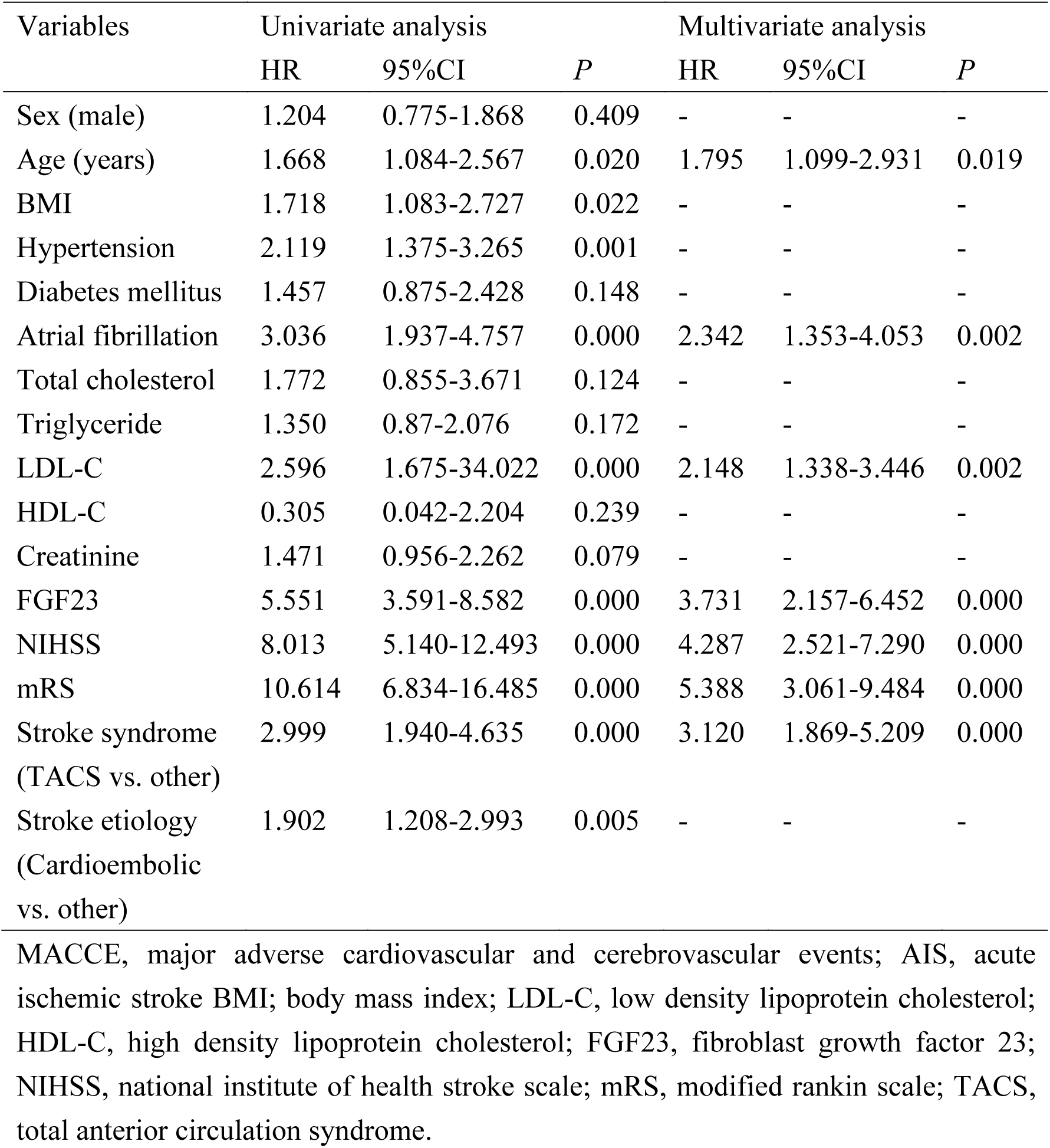
Univariate and Multivariate COX Analysis for MACCE in AIS Patients Variables Univariate analysis Multivariate analysis.

## DISCUSSION

In the present study, serum FGF23 levels were found to be elevated in AIS patients. After a median follow-up of 43 months, serum FGF23 levels were further elevated in AIS patients who incurred MACCE. FGF23 had similar high predictive value as NIHSS and mRS for the occurrence of MACCE in AIS patients. Moreover, elevated baseline serum FGF23 level was an effective predictor of the occurrence of MACCE in AIS patients.

It has been found that multiple factors involved in the development of AIS such as inflammatory response, hypoxia, energy and metabolism promote FGF23 production.^29,30^ FGF23, secreted by osteoblasts and osteoclasts, acts via endocrine forms in several organs throughout the body, including the kidneys, parathyroid glands, heart, bones and brain.^14,31^ FGF23 was found to not only inhibit the expression of NPT2a and NPT2c sodium-phosphate cotransporters, but also reduce 1,25(OH)_2_D levels to inhibit renal proximal tubular and intestinal phosphate reabsorption and thereby reduce serum phosphate levels.^32^

As a key molecule in cellular metabolic processes, phosphate plays an important role in the development of AIS by participating in energy metabolism, cellular signal transduction, inflammatory response, oxidative stress, and calcium overload.^33–35^ Reduced serum phosphate levels were strongly associated with AIS. A follow-up of 3,437 patients undergoing haemodialysis for a median of 3.9 years found that the lower the serum phosphate level, the higher the risk of AIS.^36^ As for AIS patients, reduced serum phosphate levels were associated not only with severity but also with the occurrence of MACCE.^37–39^ Inhibition of FGF23 overactivity in certain diseases caused by hypophosphatemia is considered a new therapy for the treatment of these diseases, as human MAB (burosumab) for FGF23 has been approved in several countries including Europe, North America and Japan.^32^

Elevated FGF23 were found to predispose to vascular calcification and were associated with vascular stiffness and endothelial dysfunction, all of which may contribute to AIS.^40^ The FGF23 receptor Klotho, which is widely expressed in the brain, has anti-inflammatory and antioxidant effects, and its levels decline with age and cardiovascular disease.^41^ Statins and renin-angiotensin system inhibitors, which are widely used in clinical practice, can increase Klotho levels and improve the prognosis of cardiovascular diseases.^31^

It has been shown that elevated serum FGF23 levels associated with an increased risk of AIS in individuals without chronic kidney disease.^21^ Furthermore, for these individuals, FGF23 was not only independently associated with insulin resistance, inflammation, and obesity status, but has also been shown to be a strong predictor of MACCE in patients with cardiovascular disease.^42–44^ In addition, a large number of studies found that NIHSS and mRS were predictors of the occurrence of MACCE in AIS patients s.^28,45^ This is similar to our findings that FGF23 had the same value as NIHSS and mRS for predicting the occurrence of MACCE in patients with AIS.

However, the MESA and REGARDS studies found that serum FGF23 levels were not associated with AIS or were only associated with cardioembolic stroke.^24,25^ The reason for the inconsistent findings may be related to the different study populations. For example, the MESA study enrolled people without known cardiovascular disease, whereas the REGARDS study enrolled black and white US adults ≥ 45 years of age, irrespective of cardiovascular disease. In contrast, the present study enrolled patients with AIS.

There were some limitations to this study. Firstly, this study was a single-centre study with patient enrolment in only one hospital in one region, which inevitably resulted in selection bias. Second, this study only tested the baseline serum FGF23 levels of the patients and failed to re-test their levels at the time of MACCE, which did not allow for a before-and-after controlled study. Therefore, multicentre, large-scale, long-term clinical studies are still needed to explore the relationship between FGF23 and AIS.

## CONCLUSIONS

The present study demonstrated that serum FGF23 levels were elevated in patients with AIS. Baseline serum FGF23 levels exhibited a high predictive value for MACCE in AIS patients. Furthermore, elevated baseline serum FGF23 levels were identified as a robust and effective predictor of MACCE in AIS patients.

## Data Availability

The data supporting the findings of this study are availablefrom the corresponding author upon reasonable request.

